# Cerebral blood flow pattern in patients with carotid artery stenosis with low trans-stenotic blood flow

**DOI:** 10.1101/2023.03.01.23286660

**Authors:** Laleh Zarrinkoob, Sanne Myrnäs, Anders Wåhlin, Anders Eklund, Jan Malm

## Abstract

**Background:** Compromised cerebral blood flow has been identified as a contributing risk factor for future ischemic events in patients with symptomatic carotid artery disease. Nevertheless, the hemodynamic impact of carotid stenosis is rarely evaluated. The aim of this observational cross-section study was to investigate how a reduced internal carotid artery (ICA) blood flow rate (BFR), rather than stenosis degree, relates to BFR in the cerebral arteries.

**Methods:** Four-dimensional phase-contrast magnetic resonance imaging (4D-PCMRI) was used to measure cerebral blood flow in 38 patients with symptomatic carotid stenosis (≥50%), being considered for carotid endarterectomy. The BFR in the cerebral arteries was compared between two subgroups that were based on the BFR in the symptomatic (ipsilateral) ICA: I. reduced ICA flow (<160 mL/min) i.e., 2 SD <normal ICA BFR; II. preserved ICA flow (≥160 mL/min). Furthermore, BFR laterality was defined as a difference in the paired ipsilateral-contralateral arteries within the groups.

**Results:** The degree of stenosis was not significantly different between the two subgroups (72% vs. 80%; *P*=0.09). In the reduced group compared to the preserved ICA flow group, a reduced BFR was found in the ipsilateral middle cerebral artery (MCA) (108±32 vs. 136±24 mL/min; *P*<0.01) and anterior cerebral artery (A1 segment) (−6±47 vs. 81±27 mL/min; *P*<0.001), while the BFR was increased in the contralateral A1 segment (152±79 vs. 82±41 mL/min; *P*<0.001). There was also a reversed BFR in the posterior communicating artery and ophthalmic artery on the ipsilateral side in the group with reduced ICA flow. BFR laterality was observed in all paired arteries in the reduced ICA flow group (*P*<0.05), while there was no laterality in the preserved ICA flow group (*P*>0.05).

**Conclusions:** 4D-PCMRI revealed compromised cerebral blood flow in patients with carotid stenosis, not possible to detect by solely analyzing the stenosis degree. In patients with reduced ICA flow, collateral recruitment was not sufficient to maintain symmetrical BFR distribution to the two hemispheres.

## INTRODUCTION

Patients with symptomatic carotid stenosis are at increased risk for subsequent stroke and are therefore considered for preventative revascularization.(*1*) According to current guidelines, risk stratification of these patients is based on the grade of stenosis, which is regarded to be the greatest risk factor. (*2, 3*) However, with a relatively high number needed for treatment, a more comprehensive risk stratification is desirable.(*2*) Trials have also indicated that intervention in appropriately selected asymptomatic patients results in a reduced risk for future stroke.(*4-6*) This further enhances the need for additional methods of evaluation in order to improve the precision for selecting patients where intervention may be beneficial.

Hemodynamic compromise has been identified as a contributing risk factor for future ischemic stroke.(*7*) While thromboembolism is considered the predominant mechanism of infarction due to carotid stenosis, ischemia can also be caused by hypoperfusion. Additionally, the reduced blood flow might result in impaired clearance of emboli (washout).(*8-10*) The hemodynamic state of the brain in these patients is largely dependent on collateral recruitment, which is mainly obtained through primary collateral routes of the circle of Willis and secondarily obtained through subsidiary collaterals, including the ophthalmic artery and leptomeningeal vessels.(*11*) Because of collateral recruitment, hypoperfusion does not necessarily correspond to the grade of stenosis. Additionally, signs of impairment, such as secondary collateral pathways and other known compensatory mechanisms, have been independently associated with an increased risk of ischemic events. (*7, 12-17*) Although much is known about impaired cerebral hemodynamics in patients with carotid stenosis and the associated risk of future stroke, hemodynamic assessment is rarely considered when selecting patients for intervention.(*2*)

Four-dimensional phase-contrast magnetic resonance imaging (4D-PCMRI) is a technique that allows measurement of the blood flow rate (BFR) in major cerebral arteries individually, simultaneously and noninvasively.(*18*) In a previous study using this technique, a correlation was found between stenosis degree and BFR in the internal carotid artery (ICA) but not in the middle cerebral artery (MCA), indicating that intracranial flow is maintained through collaterals.(*14*) This has also been demonstrated in the past using perfusion techniques.(*19*)

We have previously shown, using 4D-PCMRI, that due to collateral recruitment, a laterality in cerebral arterial BFR could not be explained solely by stenosis degree.(*20*) Therefore, we hypothesized that by determining the ICA BFR rather than the stenosis degree, we might gain a better understanding of cerebral hemodynamics in these patients. At length, this might help further evaluate the potential of an evolving method for hemodynamic assessment that could possibly be integrated into risk stratification of patients with carotid stenosis. The aim of the current study was to evaluate how the BFR in the ICA relates to the BFR in cerebral arteries in patients with carotid artery stenosis.

## METHODS

### Patients

Within a larger project, at the University of Umeå from 2012-2015, we prospectively recruited consecutive patients being considered for carotid endarterectomy. (*21*) Patients were eligible if they had a recent ischemic stroke or transient ischemic attack and a corresponding stenosis of at least 50%. The inclusion criteria were modified Rankin Scale (mRS)(*22*) <3 and Mini Mental State Exam (MMSE)(*23*) >23 points. The exclusion criteria were severe aphasia, contralateral carotid occlusion, atrial fibrillation, previous ischemic events, or other diseases affecting the central nervous system and contraindications to MRI. A total of 38 patients were recruited with a mean age of 72 years. Twelve patients had bilateral stenosis ≥50%, the contralateral stenoses were nonsymptomatic. The patient characteristics are shown in Table 1. The participants gave their written consent after receiving oral and written information about the procedures of the study. Approval was obtained by The Ethical Review Board of Umeå University, DNR 2011-440-31 M, and the study was performed in accordance with the guidelines of the Declaration of Helsinki.

**Table 1.** Patient Characteristics. BMI indicates body mass index; BP, blood pressure; F, female; ICA, internal carotid artery; M male; MMSE, Mini-Mental State Exam; MRI, magnetic resonance imaging; mRS, modified Ranking Scale; NIHSS, National Institutes of Health Stroke Scale; SD, standard deviation; TIA, transient ischemic attack.

| **Patient Characteristics (n=38)** |  |
| --- | --- |
| Age, years (range) | 72 (58-80) |
| Sex, M/F | 27/11 |
| Hypertension, % | 79 |
| Hyperlipidemia, % | 55 |
| Diabetes Mellitus, % | 24 |
| Previous TIA, % | 22 |
| Ever smoker, % | 72 |
| BMI, kg/m<sup>2</sup> ± SD | 27±4 |
| Systolic BP, mmHg ± SD | 138±17 |
| Diastolic BP, mmHg ± SD | 71±10 |
| NIHSS, (range) | 0 (0-6) |
| mRS, (range) | 0 (0-2) |
| MMSE, points ± SD | 27±2 |
| Subacute ischemic lesion on MRI, % | 47 |
| Ipsilateral ICA stenosis, mean % ± SD | 76±14 |
| Contralateral ICA stenosis, mean % ± SD | 41±26 |
| Bilateral stenosis ≥50%, % | 31 |

### Grading of stenosis

Thirty-one patients underwent computed tomography angiography (CTA), and stenosis was graded according to the NASCET(*24*). Seven patients had contraindications to intravenous contrast and were thus examined by carotid Doppler ultrasound. The highest peak velocity in the stenosis was translated to a stenosis grade.(*25*)

### MRI examination

Blood flow measurements for the cerebral arteries were obtained with a 3 Tesla MRI scanner (GE Discovery MR 750, Waukesha, WI, USA) using 4D-PCMRI. The median time from symptom onset to MRI examination was 6 days (range 2-60 days). The scanning time was approximately 9 minutes. For each patient, blood flow velocities were obtained for all major cerebral arteries, i.e., 19 locations. The technique has been previously described.(*21*)

### Imaging analysis and blood flow assessment

BFR measurements, in milliliters per minute, were calculated in MATLAB (The Mathworks, Natick, MA) using software developed in-house.(*26*) BFR was calculated by two investigators, LZ and AW. The images were deidentified, and the investigators were blinded to the side and degree of stenosis as well as the age, sex and medical history of the patient. BFR was measured at the locations demonstrated in *Figure 1*. Ophthalmic arteries (OAs), which are not featured, were measured close to their origin from the ICA. Total cerebral blood flow (tCBF) was obtained by adding the BFR of the ICAs and VAs on both sides. Hemisphere blood supply was defined as the sum of the BFR in the MCA, A2 segment of the anterior cerebral artery (ACA2) and P2 segment of the posterior cerebral artery (PCA2) on the corresponding side. Invisible or hypoplastic arteries were set as 0 mL/min blood flow. For the ICA, MCA, A1 segment of the anterior cerebral artery (ACA1), P1 segment of the posterior cerebral artery (PCA1) and PCA2, the mean BFR calculated by the two investigators was used. If there was a difference of 20% or more, the two investigators remeasured the BFR in that artery collectively. For the posterior communicating artery (PCoA), OA and ACA2, the two investigators measured the BFR collectively. Because the ACA2 on the left and right are too close together, measurement of the BFR in those arteries was performed using manual segmentation.

**Figure 1.**
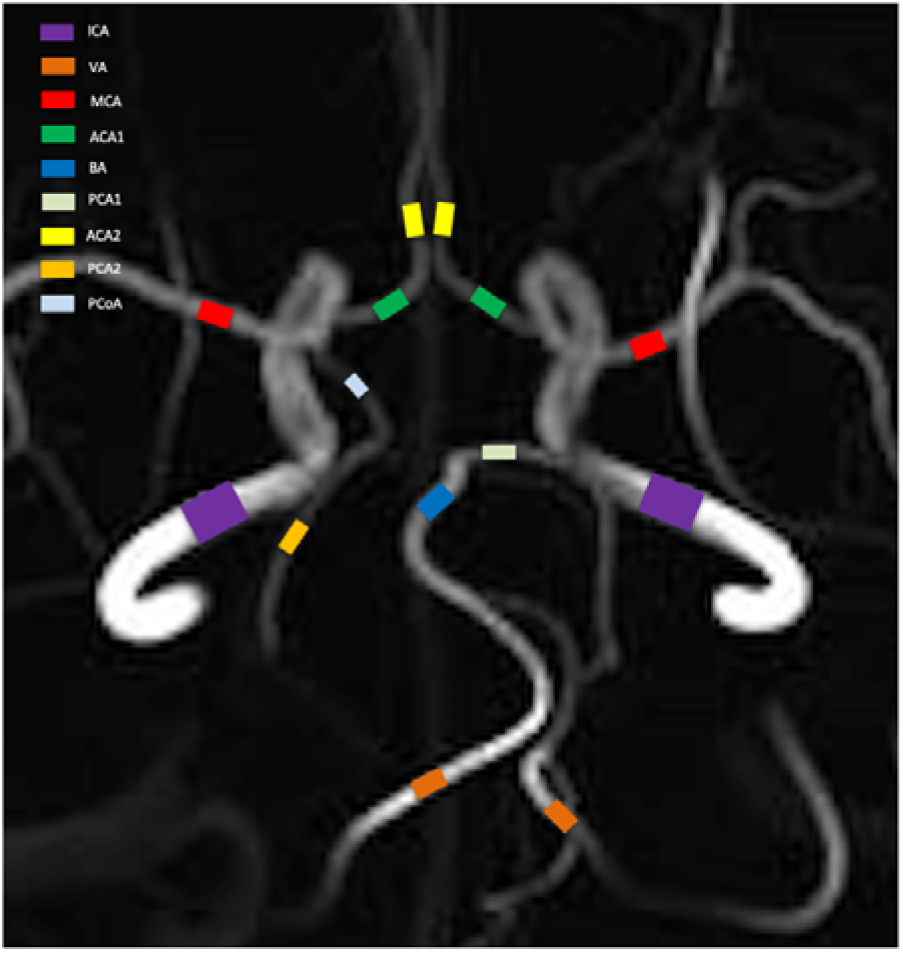
Locations for BFR measurements. ICAs, internal carotid arteries, were measured at the level of the C3-C4 segment: VAs, vertebral arteries, intracranially at the level of the V4 segment: BA, basilar artery, below the superior cerebellar artery. MCA, middle cerebral arteries, was measured at the M1 level; A1 segment of anterior communicating artery (ACA1), proximally to the anterior communicating artery and A2 segment (ACA2), distally to the anterior communicating artery: PCAs, posterior cerebral arteries, proximally (PCA1) and distally (PCA2) to the aperture of PCoA, posterior communicating artery.

### Definition of low blood flow rate in the internal carotid artery

To estimate the normal value for ICA blood flow, the PubMed Database was searched to identify trials that measured cerebral blood flow using 2D- or 4D-PCMRI in healthy elderly subjects. After initial screening, four studies were identified that presented values for ICA BFR. One study was excluded because not all groups of included subjects consisted of healthy subjects.(*27*) One study was excluded because the subjects in the appropriate age category only had a sample size of n=4.(*28*) Two studies with a total of 118 patients aged 61-80 were considered relevant.(*29, 30*) A pooled estimate for a normal ICA BFR was calculated to be 238 ± 39 mL/min. The threshold for ICA BFR was set to 160 mL/min, two standard deviations below normal.

The study population in our study was dichotomized into two subgroups based on the BFR in the ipsilateral ICA to stenosis: ICA reduced flow (n=20) with an ICA BFR <160 mL/min and ICA preserved flow (n=18) with an ICA BFR ≥160 mL/min. BFR.

### Statistical analysis

A paired *t* test was used to compare the mean BFR between the ipsilateral (carotid stenosis) side and the contralateral side. The patients were dichotomized into 2 subgroups based on ICA reduced flow and ICA preserved flow. An independent sample *t* test was used to compare the mean BFR between the two subgroups. The flow in an artery was set as 0 mL/min if the artery was either invisible or hypoplastic and its blood flow was not measurable. Values are expressed as the mean±SD, and the statistical significance threshold was set as *P*<0.05. The data were analyzed using SPSS statistics, version 27 (IBM, Chicago, IL).

## RESULTS

### Arterial inflow to the brain

There was no difference in tCBF between the group with reduced ICA flow (n=20) and the group with preserved ICA flow (n=18) (p=0.11) (Table 2). When comparing the contribution to tCBF between the two groups, ipsilateral ICA contributed to 13±12% in the group with reduced ICA flow as opposed to 35±6% in the group with preserved ICA flow (p<0.001). This was compensated in the group with reduced ICA flow by a contribution of the contralateral ICA, which was higher compared to the group with preserved ICA flow (53±13% vs. 37±7%; p<0.001). Correspondingly, the posterior supply collectively explained 34±12% and 27±8% of tCBF in the two groups, respectively.

**Table 2.** Mean BFR in cerebral arteries. **Comparison between the two subgroups based on reduced or preserved BFR**. ACA1, A1 segment of the anterior cerebral artery; ACA2, A2 segment of the anterior cerebral artery; BA, basilar artery; BFR, blood flow rate; contra, contralateral; ICA, internal carotid artery; ipsi, ipsilateral; MCA, middle cerebral artery; NA, not applicable; OA, ophthalmic artery; PCA1, P1 segment of the posterior cerebral artery; PCA2, P2 segment of the posterior cerebral artery; PCoA, posterior communicating artery; tCBF, total cerebral blood flow. * indicates P<0.05, † by definition of the subgroups.

| Arteries | ICA reduced flow<br>($\leq 160$ mL/min) | | ICA preserved flow<br>($> 160$ mL/min) | | ICA reduced flow/<br>ICA preserved flow<br>*P* value |
| --- | --- | --- | --- | --- | --- |
| | BFR mean ± SD<br>(mL/min) | ipsi/contra<br>*P* value | BFR, mean ± SD<br>(mL/min) | ipsi/contra<br>*P* value | |
| tCBF | 543 ± 116 | NA | 595 ± 77 | NA | 0.11 |
| ICA ipsi | 67 ± 60 | <0.001* | 208 ± 34 | 0.26 | <0.001* † |
| ICA contra | 294 ± 113 | | 224 ± 51 | | 0.02* |
| BA | 169 ± 62 | NA | 142 ± 51 | NA | 0.15 |
| ACA1 ipsi | -6 ± 47 | <0.001* | 81 ± 27 | 0.93 | <0.001* |
| ACA1 contra | 152 ± 79 | | 82 ± 41 | | <0.001* |
| PCA1 ipsi | 89 ± 56 | <0.01* | 56 ± 32 | 0.55 | <0.05 |
| PCA1 contra | 45 ± 39 | | 52 ± 26 | | 0.49 |
| PCoA ipsi | -11 ± 27 | 0.04* | 2 ± 29 | 0.51 | 0.15 |
| PCoA contra | 13 ± 35 | | 7 ± 18 | | 0.51 |
| OA ipsi | -7 ± 14 | <0.01* | 3 ± 8 | 0.14 | <0.01* |
| OA contra | 6 ± 7 | | 5 ± 6 | | 0.37 |
| MCA ipsi | 108 ± 32 | <0.001* | 136 ± 24 | 0.65 | <0.01* |
| MCA contra | 141 ± 41 | | 138 ± 29 | | 0.78 |
| ACA2 ipsi | 54 ± 20 | 0.01* | 59 ± 17 | 0.57 | 0.22 |
| ACA2 contra | 62 ± 22 | | 61 ± 19 | | 0.42 |
| PCA2 ipsi | 67 ± 31 | 0.04* | 56 ± 15 | 0.43 | 0.15 |
| PCA2 contra | 53 ± 23 | | 58 ± 14 | | 0.47 |

### Flow in the Circle of Willis

In summary, the difference in collateral circulation between the two groups was most pronounced in the ACA1s, where the BFR was reversed ipsilaterally and increased contralaterally in the group with reduced ICA flow. For the posterior collaterals (PCA1 and PCoA), there was an augmented BFR (reversed in PCoA) toward the ipsilateral side in the group with reduced ICA flow. The OA BFR was also reversed in this group (Table 2). In the group with reduced ICA flow, a laterality was found in all cerebral arteries when comparing the BFR in ipsilateral and contralateral arteries, while there was no BFR laterality in the group with preserved ICA flow (Table 2 and Figure 2).

**Figure 2.**
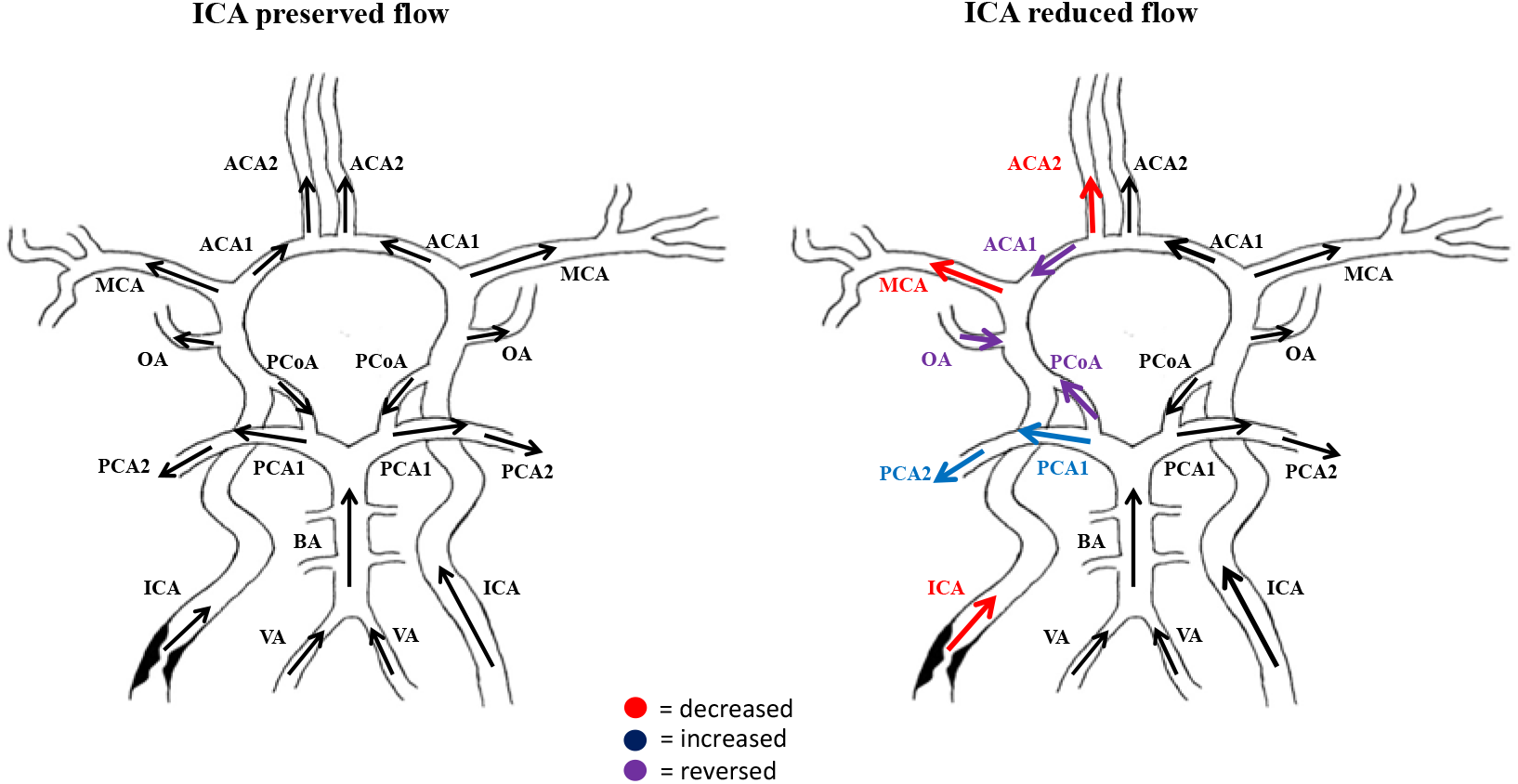
Comparison between the ipsilateral (left) and contralateral (right) sides in the two subgroups. Red indicates decreased BFR, purple indicates reversed BFR, and blue indicates increased BFR. Abbreviations: ACA1, A1 segment of the anterior cerebral artery; ACA2, A2 segment of the anterior cerebral artery; BA, basilar artery; ICA, internal carotid artery; MCA, middle cerebral artery; OA, ophthalmic artery; PCA1, P1 segment of the posterior cerebral artery; PCA2, P2 segment of the posterior cerebral artery; PCoA, posterior communicating artery.

### Hemispheric blood supply

Despite evident collateral recruitment in the group with reduced ICA flow, the BFR in the ipsilateral MCA was still significantly lower compared to the contralateral side and compared to the group with preserved ICA flow (Table 2). Notwithstanding the compensatory increase in the ipsilateral PCA2 BFR, the total supply of the hemisphere was still significantly lower than that on the contralateral side (on the ipsilateral (228 ± 53 mL/min vs. 256 ± 65 mL/min; *P*<0.01). No laterality was evident in the group with preserved ICA flow for individual arteries (Table 2, Figure 2) or for hemispheric blood supply (*P*=0.29). The laterality in the hemispheric blood supply is illustrated in *Figure 3*.

**Figure 3.**
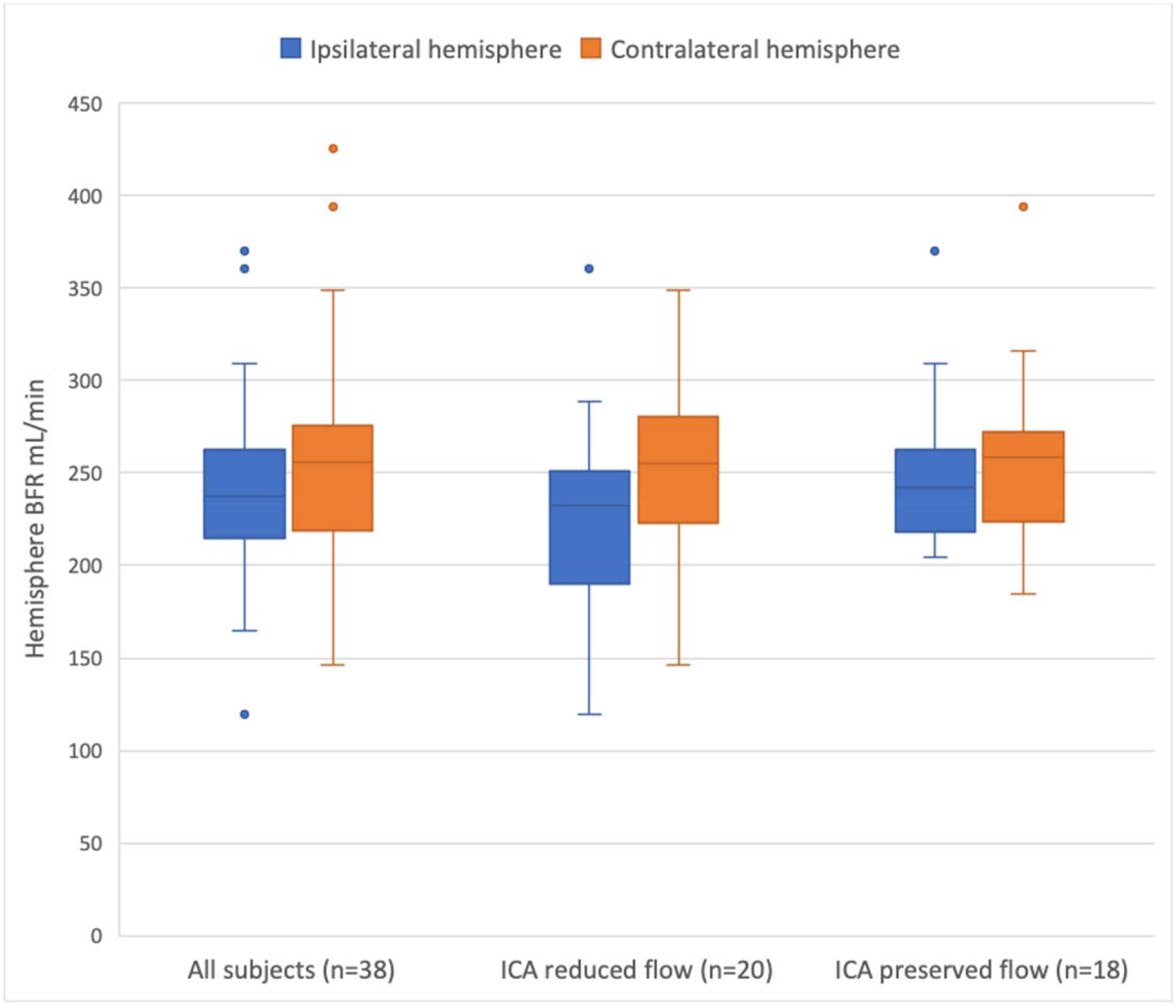
Ipsilateral and contralateral hemisphere blood supply. Hemispheric blood supply (ACA2, MCA and PCA2 BFR) in the entire cohort as well as within the two subgroups. A significant difference (*p*=<0.01) between ipsilateral and contralateral hemisphere blood supply was seen in patients with reduced ICA flow. BFR indicates blood flow rate; ICA, internal carotid artery.

### Characteristics and vascular features of the two subgroups

None of the characteristics described in Table 1 differed significantly between the two subgroups. The mean stenosis degree in the symptomatic ICA was 80% in the group with reduced ICA flow and 72% in the group with preserved ICA flow (*P*=0.09). In the group with reduced ICA flow, 35% had bilateral stenosis, and the mean degree of contralateral ICA stenosis was 46%. In the group with preserved ICA flow, 28% had bilateral stenosis, and the mean degree of contralateral ICA stenosis was 36%.

## DISCUSSION

Our study has shown that distal cerebral hemodynamics can be markedly different in subgroups of patients with similar stenosis grade but different ICA blood flow. PCMRI can therefore be a tool for hemodynamic assessment that could possibly be integrated into preoperative risk stratification of patients with carotid stenosis.

### 4D-PCMRI and possible clinical applications

4D-PCMRI allows for direct measurement of BFR in the cerebral arteries simultaneously and noninvasively. While perfusion methods are only able to look at the end result, with 4D-PCMRI, one can assess the blood supply to different regions of the hemispheres and study the routes by which the blood supply gets there.

In patients with carotid artery disease, possible clinical applications of the technique consist of the opportunity to select patients more carefully with a potential benefit by intervention. This might be particularly relevant in patients with asymptomatic stenosis. Signs of hemodynamic impairment along with severe stenosis have been associated with an increased risk of future ischemic stroke, independent of symptomatic presentation.(*12*) Furthermore, in a recent cohort of asymptomatic patients, improved hemodynamics following carotid revascularization was associated with improved cognitive function.(*32*) Some of the patients with reduced ICA flow in this study are likely patients with near-occlusion, a diagnosis that is often overlooked in routine practice and where the recommended treatment differs.(*33-35*) Our results indicate that in these patients, the reduced ICA flow affects the flow in more distal cerebral arteries.

### Inadequate collateral compensation in patients with abnormally low ICA flow

We have previously investigated how the degree of stenosis relates to the cerebral arterial BFR. We failed to show that severe stenosis (≥ 70%) was associated with reduced flow in the ipsilateral MCA.(*20*) Previous studies have also demonstrated that the severity of stenosis is not the sole determinant for ICA blood flow as residual lumen and stenosis length are also involved.(*14, 36*) Furthermore, it is well recognized that the severity of stenosis alone cannot predict the hemodynamic impact of carotid artery disease because of collateral recruitment.(*19*) With our study, we were able to demonstrate that on a group level, patients with reduced ICA flow are also subject to hemodynamic changes in more distal arteries, independent of the severity of the stenosis. The reduced MCA flow could possibly increase the risk of future ischemic events due to impaired clearance of emboli and vulnerability to hypoxia and fluctuations in blood pressure.(*7, 8*)

The finding of reduced blood flow in the ipsilateral MCA in the group with reduced ICA flow suggests that in these patients, collateral flow did not provide sufficient compensation. The detected BFR reduction was inspite of a potentially reduced vascular resistance by the autoregulatory vasodilation. However, the presence and degree of autoregulatory vasodilation were not assessed in the patients in this study.

Another possible explanation for a reduction in MCA flow is decreased metabolic demand as a result of neuronal injury. Although this was not accounted for in this study, there was no difference between the two subgroups regarding the presence of ischemic lesions on MRI, nor was there a difference in the MCA BFR between the patients with or without ischemic lesions on MRI (*Table 1*). In addition, we have previously shown that a low BFR in the MCA, seen in a subgroup of patients with collateral recruitment before carotid endarterectomy (CEA), was normalized after CEA. This finding pointed toward a hemodynamic disturbance in the MCA territory as the main reason for the low MCA BFR in this group of patients.(*37*)

### Collateral recruitment and laterality in hemisphere blood supply

When comparing the two groups, it is evident that in the group with reduced ICA flow, collateral flow is mainly derived from the contralateral ICA through the anterior arteries of the circle of Willis. As previously suggested, it seems that collateral flow via this route supplies the ACA2 territory bilaterally.(*21*) However, when comparing the ipsilateral and contralateral sides, blood flow in the ipsilateral ACA2 is still decreased, indicating no further compensation via ACA2. In contrast, the augmentation of flow via posterior collaterals toward the ipsilateral side resulted in an increased BFR in ipsilateral PCA2, which might indicate compensation via leptomeningeal collaterals.(*38*) Additionally, in patients with reduced ICA flow, this collateral recruitment was not sufficient to provide a symmetrical distribution to the two hemispheres. Collateral flow from the contralateral ICA could reasonably be impeded by the presence of a stenosis on that side as well. However, the two subgroups did not differ regarding the presence or degree of contralateral stenosis.

### Limitations

The limitations of this study are its small sample size and the fact that the included patients were heterogeneous in regard to their ipsilateral and contralateral stenosis. However, the study population consisted of consecutive patients considered for carotid endarterectomy and is therefore representative of routine clinical practice.

With 4D-PCMRI, there are some method-specific problems with distorting noise. It is likely that very low flow velocities cannot be distinguished from noise, and the complete absence of flow could not be firmly established. 4D-PCMRI can also be distorted in the region of the orbit, making it more difficult to measure flow in the OA.

Finally, a limitation of this study is the lack of a control group to determine normal ICA blood flow. By using data from previous trials, however, normal ICA blood flow could be estimated based on a larger population.

## Conclusion

Reduced ICA flow was associated with reduced BFR in the ipsilateral MCA, irrespective of the stenosis grade. Collateral recruitment was evident: the anterior collaterals supplied the ACAs bilaterally, and there was an augmentation of flow toward the ipsilateral side via the posterior collaterals. However, collateral recruitment was not sufficient to maintain normal blood flow to the MCA territory, nor did it provide a symmetrical distribution to the two hemispheres. Thus, 4D-PCMRI revealed compromised cerebral blood flow in patients with carotid stenosis, which was not possible to detect by solely analyzing the stenosis degree. In patients with reduced ICA flow, collateral recruitment was not sufficient to maintain symmetrical BFR distribution to the two hemispheres. It is plausible that, with further study, this technique might be used to improve risk stratification for these patients.

## Data Availability

All data referred to in tis manuscript is available on request

## Sources of Funding

The following financial support has been conducted for the research, authorship, and publication of this article: the Swedish Research Council, grant numbers 2015–05616; 2017– 04949; the County Council of Västerbotten; and the Swedish Heart and Lung Foundation, grant number 20140592.

## DISCLOSURES

There are no conflicts of interest to report.

## Notes

**Disclosures:** No conflict of interest to report.

### Competing Interest Statement

The authors have declared no competing interest.

### Clinical Trial

The study was not interventional and did not evaluate any drugs.

### Author Declarations

Approval was obtained by The Ethical Review Board of Umeå University, DNR 2011-440-31 M, and the study was performed in accordance with the guidelines of the Declaration of Helsinki.

